# The Dementia Trials Accelerator (DTA): a UK dementia trials-ready cohort

**DOI:** 10.64898/2026.08.14.26359869

**Authors:** William Whiteley, Cornelia M. van Duijn, Neil Postlethwaite, Emily Beal, Kimberly Bennett, Graham Blakoe, Helen Brooks, Katharine Collet, Paul Elliott, Eoghan Forde, Amanda Heslegrave, Lorraine Holland, Ivan Koychev, Jo Latimer, Thomas Littlejohns, Paresh Malhotra, Matthew Retford, Katie Smith, Jonathan M. Schott, Amy Tilbrook, Joanne Thomas, Rhoswyn Walker, Helen Ward, Henrik Zetterberg, Monika Ziminska, Andrew Morris, Siddharthan Chandran

## Abstract

The Dementia Trials Accelerator (DTA) is a UK-wide programme designed to improve the feasibility, efficiency, and inclusiveness of recruitment into clinical trials of dementia and related brain-health conditions. Dementia trials are frequently constrained by the difficulty and cost of identifying eligible participants, which often requires cognitive assessments and measurement of blood-based biomarkers.

The DTA addresses these barriers with two linked services.

First, the DTA provides a federated platform to improve findability of potential participants across existing UK-based cohorts with consent to recontact. A single point of contact across multiple cohorts would allow increased efficiency of search for participants for studies.

Second, the DTA provides a community-centred pre-screening service with information relevant to trial eligibility and a linked plasma and DNA tissue bank, with participants’ consent for recontact. Participants aged 65-75 years are approached via existing cohorts and registries. Consenting participants complete an online questionnaire and digital cognitive assessment, attend an in-person assessment for physical measures and face-to-face cognitive testing, and provide venous blood samples which are processed to plasma and whole blood for long-term storage and biomarker measurement. The initial programme target is to recruit at least 10,000 participants into the DTA pre-screening service.

The DTA is designed to support approved academic and industry studies by enabling the DTA team to approach eligible participants for specific studies, without transferring identifiable information without consent. In parallel with its immediate trial-readiness purpose, the DTA is positioned to interoperate with emerging national approaches to biomarker-led recruitment by generating high-quality, recontactable cohorts with standardised cognitive characterisation and scalable biosampling suitable for blood-based biomarkers.

## Introduction

Randomised controlled trials (RCTs) are a critical bottleneck in the development of new agents for the treatment and prevention of dementia. In 2025, there were 182 RCTs of agents for dementia, and delivering these trials as fast as possible is a priority: a delay of one day in clinical trial delivery is estimated to cost a sponsor of neurology trials about $40,000 (range $13,000 to $92,000).^1,2^ Taking part in, or delivering, these RCTs give participants the opportunities to try innovative agents, clinicians important experience with new models of delivery, and the health systems financial benefits, particularly where commercially sponsored trials bring research income.^3^

However, recruitment to RCTs to prevent or treat dementia is a major challenge in any health system. Although people with dementia and their families are often highly motivated to take part in research, many cannot if they live far from centres of excellence or if they have not learned about RCTs from clinical services. Once they have volunteered to join a study, many people do not meet precise RCT inclusion criteria - they are ‘screened out’. This is frustrating to potential participants and costly to sponsors, because screening is time consuming and expensive.

Therefore, health systems need to improve accessibility to RCTs, shorten time to recruitment, and reduce screen failures.

Participants often take part in RCTs after direct invitation from a clinician. However, strained clinical services cannot approach all potentially eligible participants with symptoms of cognitive impairment, which means many people miss the opportunity to take part. For RCTs focused on dementia prevention, eligible people without symptoms may never even encounter dementia services. Widening access pathways would shift decision-making power from clinicians to potential participants themselves, which could lead to volunteers taking part in studies that they would otherwise never have heard about.

In the UK, up to 1 in 30 people take part in cohort studies, and more participate in National Institute of Health and Care Research (NIHR) registers, such as Join Dementia Research (JDR) and Be Part of Research.^4,5^ These people, who have already volunteered to take part in studies, have contributed an enormous amount to UK research and could contribute further to future RCTs in dementia treatment and prevention. Programs exist in the UK to search for participants across cohorts for new RCTs,^6^ although there is scope to strengthen co-ordination between cohorts and participation in trials.

Therefore, we developed the Dementia Trials Accelerator (DTA), a service to recontact participants in UK-based cohorts and registers for trials in dementia and brain health; to deliver remote screening for RCT eligibility, with cognitive testing and face-to-face blood draw for blood-based biomarker measurement; and to develop an interface with clinical trial sites. This combination of services has been recommended by policy makers^7^ and delivered to varying degrees across the US^8–11^ and the UK^12–15^. In addition, it aligns with UK government strategies to improve the use of existing data resources for clinical trials and bring RCTs closer to the homes of potential participants.^3^ Here we describe the design of the DTA services: a federated search capability across UK cohorts for participants with consent for recontact; enhanced phenotyping of potential participants; and recontact with participants for RCTs.

### The DTA cohort search platform

The DTA used cohort discovery technology developed by Health Data Research UK (HDR UK) to allow researchers to specify participant criteria, query multiple cohorts, and deliver counts of people with particular characteristics. Data itself never leaves its source: queries are federated to cohort data held within secure data environments, with only aggregate counts returned. These counts are rounded, and small-number suppression is applied to reduce the risk of participant re-identification. The DTA has worked with a pioneer group of cohorts to map their dementia-relevant data to the Observational Medical Outcomes Partnership (OMOP) common data model in partnership with the Health Informatics Centre at the University of Dundee, in order to support federated queries.^16^ These cohorts, which all have permission to recontact participants, are: Generation Scotland,^17^ JDR,^5^ NIHR Bioresource Genes and Cognition,^18^ the REal-time Assessment of Community Transmission cohort (REACT),^19^ Great Minds,^6^ and the Scottish Health Research Register & Biobank (SHARE).^20^ Currently the DTA can only provide counts of potential participants and connect potential users with cohorts, but its ambition is to develop a service to recontact participants for RCTs through their parent cohorts.

### The DTA phenotyped cohort

To test the mechanism for recontact with cohort participants and community-based trial pre-screening, the DTA worked with REACT to recruit and phenotype a DTA trial ready cohort. The REACT study was originally designed to estimate the prevalence of the coronavirus SARS-CoV-2 across England during the COVID-19 pandemic.^21^ At sign-up, REACT had asked participants for permission to recontact them with future research opportunities (ie consent for recontact), and a large number of people (>2.5 million) had joined from across the whole of England.^19^ REACT identified potentially eligible participants, and emailed them an invitation to join the DTA. The email contained a link to a secure DTA participant signup website with information describing the purpose of DTA, procedures involved, and data handling arrangements. If interested, participants provided electronic consent and completed an online questionnaire and digital cognitive assessment. They then booked an appointment at a nearby assessment centre using a unique web link, with reminders delivered by email and text.

Eligible participants were UK-based, known to be alive at invitation and (for the initial cohort) aged between 65 years to 75 years inclusive. The age group was chosen as the most relevant for dementia prevention RCTs. Exclusion criteria were primarily practical: lack of consent, due to leave the UK, inability to travel to a clinic appointment or trial site, or that they declined to be contacted about RCTs of investigational medical products aimed at dementia prevention or treatment. We did not exclude people with a self-reported diagnosis of mild cognitive impairment, dementia, or major psychiatric or neurological illnesses, but these were recorded in the baseline questionnaire. Participants were offered vouchers in lieu of travel-cost reimbursement if travel would otherwise prevent participation. The DTA team regularly monitored sociodemographic and regional information of invited and recruited participants to identify potential under-represented populations relative to the UK population between 65 and 75. Where monitoring identified under-representation, additional recruitment strategies, including targeted invitations, or reimbursement, could be used.

A staged consent process was used. Consent was obtained electronically at registration, following provision of online participant information materials. Consent was confirmed during an in-person assessment, which included review of participant understanding and an opportunity to ask questions. DTA recruited only participants judged to have capacity to consent to participation. Capacity was assessed at the in-person visit by trained staff using standard operating procedures. Where staff had concerns that a participant did not understand the study procedures or implications of consent, participation did not proceed and appropriate support was offered. Participants may have taken part because they were concerned about dementia or had concerns about their personal or family risk. The Alzheimer’s Society was a partner to the DTA programme and provided an independent source of counselling to potential participants, participants and families. At contact with the Alzheimer’s Society, participants were guided to resources depending on their need.

Participants consented to data collection, remote and in person cognitive testing, blood draw, anthropometry, data linkage, and to recontact for clinical studies based on the information the DTA held about them.

Participants were asked to complete an online questionnaire (Table 1) after consent which typically needed 20–30 minutes to finish. The questionnaire was designed to support RCT pre-screening and included contact details, socio-demographics and occupation, cognitive function and functional ability, family history, psychiatric and medical history, and general health, including items aligned with inclusion and exclusion criteria from recent dementia RCTs.^21–24^ Participants were asked to consent to linkage of their data with NHS and other health-related records across primary and secondary care to identify health-related events relevant to study eligibility.

**Table 1.** Data collected at baseline questionnaire and at assessment.

|  |  |
| --- | --- |
| **Baseline questionnaire** |  |
| Demographics | Age, sex, gender, ethnicity, language |
| Medical history | Cardiovascular, metabolic, psychiatric, neurological diseases, sensory impairments and cancer, cardiovascular and neurological medicines, previous brain scanning and suitable for future scanning |
| Dementia and cognitive history | Mild cognitive impairment, dementia diagnosis, family history of dementia and neurodegenerative diseases |
| Activity | Physical activity, sleep, smoking, familiarity with games |
| Education and employment | Education, current employment, |
| Cognition | Cognitive function instrument questions, Cognitron battery: Object Recognition Memory Immediate, 2D Manipulations, Switching Stroop, Verbal Analogies, Word Definitions, Spatial Span, Object Recognition Memory Delayed |
| **In person visit** |  |
| Cognition | Mini-mental status examination |
| Anthropometry | Blood pressure, height, weight, |
| Blood draw | Plasma, buffy coat rich sample |
| **Biomarkers** | Glial fibrillary acidic protein, neurofilament light chain, phospho-tau-217, creatinine (where measured) |

Participants also completed a digital cognitive test (Table 1). Digital cognitive tests generally have a high acceptability, although some participants need support during testing.^25^ We convened a panel of dementia and cognitive testing experts to review existing tools. The panel agreed that a remote, digital cognitive assessment for the DTA should: take no more than 20 minutes; measure general cognition, word and visual memory; be validated in community-dwelling people with varying impairment levels; have been tested against established measures (e.g. Montreal Cognitive Assessment^26^ or Mini-Mental Status Examination (MMSE)^27^), and be deliverable via an application programming interface (API) accessible across different devices. We asked REACT participants and people affected by dementia to review a shortlist of suitable tests, who felt that all shortlisted tests were suitable. The tests that fitted our needs best were supplied by Cognitron, which was then integrated into the participant signup journey.^28^ Participants had access to email and telephone support if they needed it.

Participants attended an assessment centre where their identity was verified and consent was confirmed (<u>Table 1</u>). At the clinic visit, trained staff measured height, weight, and blood pressure with clinically validated devices. If blood pressure was above recommended thresholds for NHS community screening^29^, participants were advised to repeat blood pressure measurement at a primary care site. Participants also completed a face-to-face MMSE. Staff collected venous blood in two 9 mL K_2_EDTA Vacutainers. Blood was immediately centrifuged on site (and at latest within 2 hours of collection) at 2000 relative centrifugal force for 10 minutes, and plasma and buffy coat rich layers were aliquoted into pre-barcoded tubes and frozen at -20°C before transport via monitored cold chain to central biobank storage within seven days. On receipt, stored aliquots were transferred to -80°C freezers, with aliquots divided between the originating partner and the DTA programme for long-term storage, enabling future approved research use. DNA was extracted from whole blood and stored long term at -80°C. One aliquot of plasma was used to measure p-tau217, glial fibrillary acidic protein (GFAP) and neurofilament light chain (NfL) with the Quanterix ALZpath p-tau217 Advantage PLUS (cat no. 104570), and the Neurology 2-Plex B Advantage PLUS (cat no. 104670) kits, respectively, and creatinine using validated clinical chemistry assays.

P-tau217 is an important blood biomarker of cerebral amyloid-β (Aβ) plaque and neurofibrillary tau tangles, the pathological hallmarks of Alzheimer’s disease (AD).^22^ Higher p-tau217 can occur many years before dementia symptoms, and is associated specifically with AD pathology and amyloid accumulation.^23–25^ Using a Quanterix p-tau217 assay, in an unselected population based sample of Norwegian 65-to 74-year-olds, 23% had intermediate (0.40-0.63 pg/mL) and 16% had high (≥0.63 pg/mL) levels.^26^ Combining plasma p-tau217 with lower digital cognitive assessment scores can also identify a population at greater risk of cognitive decline over the following four years.^27^ Creatinine was measured to estimate renal function, as it has previously been demonstrated to increase p-tau217 levels possibly by reducing renal clearance, although this is still controversial.^28,29^ With this evidence, p-tau217 is increasingly a part of RCT screening protocols, particularly those aiming to recruit people for RCTs for AD prevention. GFAP, a marker of reactive astrogliosis, and NfL an axonal structural protein, were chosen as more general markers of astrocyte and neuronal damage. They are raised in neurological and neurodegenerative disorders including traumatic brain injury, stroke, multiple sclerosis, AD, and several dementias.^30^ Biomarker thresholds for trial pre-screening will be study-specific. DTA will not use a single universal p-tau217 cut-off for all studies and will be guided by RCT inclusion criteria thresholds accounting for, where appropriate, factors such as renal function.

Buffy coat rich plasma is stored for later DNA extraction and genotyping.

The DTA informed participants about clinically actionable measurements via an email with a link to a results portal, after assessment. The results shared included blood pressure, body mass index, and results from creatinine tests (where measured) with an indication of whether values outside normal ranges and advice on action to take. Participants could access their online cognitive test results if they wished, with careful explanation to put results in context. However, participants did not consent to be informed about non-actionable results, and therefore they were not told their levels of p-tau 217, GFAP or NfL. Participants could withdraw from DTA at any point, by withdrawing from further contact, access, or further use of data, and request destruction of samples (or all of these).

In order to facilitate rapid recruitment of participants, after an initial scale up phase, 8 clinic sites were actively recruiting at any one time across England for five to six days a week, with daily review of response rates, data collection, and research clinic functions. Sites were chosen to be easily accessible to participants (with easy access by car and public transport), with geographic dispersal across England, and close to NHS research sites that were actively recruiting to dementia-relevant RCTs.

### Recontact for trials

A central function of DTA is to work with approved academic and industry studies to identify and approach eligible participants for RCTs. DTA does not transfer personal details without participant consent; instead, it contacts eligible individuals and provides details on how to take part in new RCTs. DTA participants can then choose whether to take part in the studies they are offered, which prevents a one-way-street to inevitable RCT inclusion.^31^

However, recontact has ethical implications, particularly where RCT inclusion criteria include genetic or blood biomarkers of future dementia risk. Invitation to a study based on these markers could implicitly reveal that a participant belongs to a risk-stratified subgroup. However, biomarkers and cognitive measures are not deterministic. They are currently not sufficiently reliable for individual-level prediction in asymptomatic populations, but there is a risk that participants infer more than the evidence supports.^32,33^ Evidence may emerge that therapies targeted with p-tau217 or other blood biomarkers prevent dementia, although at the time that DTA was designed, there is no established therapeutic strategy for people with a raised p-tau217 but no dementia clinical syndrome. Therefore, recontact based on these levels creates a tension between respect for the right to know, and the right not to know a result that is not directly actionable.

Empirical evidence from recall-by-genotype, dementia-related *APOE* and amyloid disclosure qualitative and quantitative studies with in-person counselling does not generally support the hypothesis that revealing unknown risk factors leads to general harms. Participants do not generally regard genotype-based recontact as problematic - in Avon Longitudinal Study of Parents and Children (ALPSAC), few expressed immediate concerns and many participants favour a lower threshold for return of individual findings than professional guidance typically assumes.^34,35^ This suggests that targeted recontact is not generally inherently harmful. In dementia, psychological harms are uncommon when using structured protocols. A systematic review found little evidence of substantial harms following disclosure of Alzheimer’s disease biomarkers.^36^ Similarly, people with a family history of Alzheimer’s disease who learned they had *APOE*4 did not show increased anxiety after a 90-minute disclosure session, and participants rarely regret receiving results, and some report benefits from testing.^37^ However, knowledge of *APOE* genotype has been associated with poorer self-assessment of cognition in healthy older adults.^38^ More broadly, disclosure of elevated risk may turn otherwise healthy individuals into “patients-in-waiting,” with consequences for identity, future planning, and perceptions of health.^39^ The core question is how to recontact in a way that respects autonomy, protects welfare, preserves privacy, and supports informed participation.^39^

For the DTA, we elected to inform people about their RCT eligibility, but not the particular reasons for approach routinely. If the screening for a RCT required the measurement of a particular biomarker, this would be repeated at the clinical trial site and fed back to the participant face-to-face by the RCT team. We elected for this approach, principally because a biomarker level is not clinically actionable, feedback is likely to lead to anxiety in a minority of participants, and we did not have resources for extensive face-to-face counselling. Trial invitations will state that potential eligibility may be based on a combination of measures made by the DTA. They will also state that an invitation does not mean that the participant has dementia or will develop dementia, and that full eligibility can only be determined by the recruiting trial team.

DTA participants will take part in studies run by the UK NHS clinical research infrastructure, which is delivered through a network of trial sites based in primary and secondary care funded by the NIHR. There is a Research Delivery Network (RDN)^40^ that is condition agnostic and generally delivers phase 3 RCTs in dementia and other conditions, but nested within it are sites with particular expertise for phase 1 and phase 2 studies in dementia, the Dementia Trials Network (DTN).^41^

### Patient and public involvement and engagement (PPIE)

Public and patient perspectives were captured via working with members of public advisory and interest groups affiliated to HDR UK, Imperial and the REACT study. Public contributors provided input throughout the design of the study, including for ethics documentation, recruitment, registration, opt-outs and withdrawals, and evaluation. The DTA included an internal pilot with a mixed-methods evaluation to assess acceptability. Key metrics included participation proportions among those approached and attendance proportions among those booking appointments.

Acceptability is also evaluated with participant satisfaction surveys, reviews of data collection and complaints, clinic observation and a programme of interviews planned with participants and those who opt out or withdraw from the DTA.

### Sample size

No formal sample size calculation was undertaken. The target was to recruit individuals with and without symptoms of mild cognitive impairment and dementia, with an initial goal of 10,000 completing an assessment within the first 12 months to demonstrate feasibility of these services. We will assess feasibility of recruitment by estimating the proportion consenting among invited; and of those consenting, the proportion completing online questionnaire; digital cognitive testing; booking clinic appointment; and attending appointment. We will compare the recruited population with UK Census or ONS data for age, sex, ethnicity, deprivation, geography; and with the population in REACT. The proportion above different p-tau217 threshold will be an important metric for a population of people consenting for RCT-based screening. The ultimate test will be the proportion of people fulfilling eligibility criteria who take part in an RCT.

### Management

The DTA has NHS Health Research Authority (HRA) research ethics committee (REC) approval as a Human Tissue Authority (HTA) research tissue bank from the South-Central Berkshire B Research Ethics Committee (25/SC/0317). This means that researchers wishing to use the resource do not need separate ethics approval if recruitment through the DTA is mentioned in their own REC approval. Studies can request the DTA team to approach participants meeting their inclusion criteria, subject to review by a DTA Access Committee and appropriate approvals. DTA is managed through a multi-organisation structure. Academic and commercial studies will be considered through the same controlled access pathway. Access decisions will consider scientific validity (determine by in progress or available industry or academic funding), ethical approval (determined by progress or completed REC review), public benefit and alignment with DTA participant consent. External studies will not receive identifiable participant-level information unless a participant actively consents to contact or enrolment. It is hosted by HDR UK and the UK Dementia Research Institute, which receive funding from Office for Life Sciences through the UK Medical Research Council to deliver the DTA. Imperial College London is the establishment responsible for management of the tissue bank, and biological material is stored at a contracted laboratory facility operating under an appropriate Human Tissue Authority licence.

## Discussion

The Dementia Trials Accelerator (DTA) will bridge the gap between population cohorts, research registries, and clinical trials by lowering barriers to identifying, characterising, and approaching potentially trial-eligible participants. The model combines online assessment, remote cognitive testing, in-person clinical measurement, and high-quality bio-samples, which allows biomarker-informed pre-screening and recontact.

Participants in cohorts and research registries are willing to contribute to health research, but it is often difficult for them to take part in trials. The DTA facilitates participation in dementia trials by providing a structured route from cohort participation to trial invitation. The pre-screened cohorts of potential participants (defined either with variables in existing cohorts, or with DTA pre-screening), have advantages of faster recruitment, fewer screen failures, and ultimately lower costs for sponsors. However, how far pre-screened cohorts are better than “just-in-time” recruitment is uncertain and likely to vary by RCT design, eligibility criteria, biomarker thresholds, and geography. Evaluation of the DTA and other trial ready cohorts needs to include the number of participants recruited, invitation response rates, trial-site referral rates, screen-failure rates at trial sites, time to randomisation, participant burden, cost per randomised participant, and trial completion rates.

Risk-based recontact raises ethical issues. Invitation to a trial based on biomarkers, or other dementia risk factors may implicitly signal that a participant belongs to a higher-risk subgroup. Invitations will explain that eligibility may be based on a combination of factors, and that further assessment at a trial site is usually required. Consent is an ongoing process rather than a one-off event. Over time, participants may not remember the details of their original consent or the meaning of biomarker-informed recontact. For this reason, RCT invitations should adopt an “as-if-first-contact” approach: providing clear information, avoiding assumptions of prior understanding, and making it straightforward for participants to decline, pause, or withdraw from future contact.

Consent to recontact is central to the DTA model, but cohorts vary in governance arrangements, consent, and willingness to approach participants about RCTs. A federated approach respects cohort governance and avoids unnecessary transfer of identifiable data but also introduces potential variation in speed of participant approach. If the DTA is to deliver rapid recruitment, clear and harmonised governance is needed, ideally with transparent expectations. The DTA phenotyped cohort provides an alternative model in which the DTA itself becomes the primary research home for participants. This has operational advantages because participants consent directly to DTA processes, including future RCT contact.

If studies seek only participants with elevated p-tau217 or other specific biomarker profiles, many DTA participants without these profiles may not receive invitations and could be disappointed.

Conversely, those who fall into highly sought-after biomarker-defined groups may be approached repeatedly. The DTA will monitor the frequency and nature of recontact, to prevent overburdening particular subgroups. The research landscape should include a broad portfolio of studies, including RCTs and observational studies relevant to people with different biomarker profiles, vascular and lifestyle risk factors, cognitive trajectories, and broader brain-health outcomes.

Like most clinical research, the DTA will be affected by selection and consent bias. Participants will likely differ from the wider population in education, socioeconomic position, digital literacy, comorbidity, and willingness to engage with research. The DTA’s use of online assessment has major scalability advantages, but may also exclude people with limited internet access, low digital confidence, sensory impairment, language barriers, or cognitive difficulties. The requirement to attend an in-person blood-draw appointment may further reduce participation among people with mobility limitations, caring responsibilities, financial constraints, or poor transport access. Where under-representation is identified, during recruitment we will over-invite under-represented groups and already offer voucher in lieu of travel-cost reimbursement, and flexible appointment locations and times.

Patient and public involvement has been integral to DTA development and should continue throughout implementation. Participant input is especially important for the design of invitation materials, acceptability of risk-based recontact, return of results, withdrawal processes, and policies on repeated approach. Continued involvement will help ensure that the DTA remains aligned with participant expectations and public trust, rather than becoming solely a recruitment mechanism for sponsors. The DTA should also ensure equitable access for both academic and industry-sponsored studies, with transparent prioritisation when multiple studies seek to approach the same participant groups.

The DTA provides scalable UK infrastructure for identifying, phenotyping, and recontacting potential participants for dementia and brain-health trials. By combining federated cohort discovery, standardised cognitive and health assessment, blood-based biomarkers, and participant recontact, DTA aims to reduce recruitment delays and screen failures while widening access to trials. Sponsors and investigators of RCTs and clinical studies are invited to contact the DTA team to use the DTA services.

### Profile in a nutshell

- The Dementia Trials Accelerator (DTA) is a UK-wide pre-screening platform and linked plasma/DNA tissue bank and database established to support more efficient, scalable recruitment into clinical trials of dementia and other brain-health disorders.
- DTA recruits adults aged 65–75 years from existing cohorts/registries and NHS-linked routes (initially including collaboration pathways with cohorts such as REACT and registries such as Join Dementia Research), focusing on people at elevated dementia risk primarily by virtue of age and other trial-relevant characteristics.
- Participants undertake an online questionnaire and digital cognitive assessment, followed by an in-person assessment that includes physical measures (height, weight, blood pressure) a face-to-face cognitive assessment, and venous blood sampling (approximately 20 mL).
- Blood samples are processed to generate plasma and whole blood for DNA extraction, stored long-term (with centralised logistics and monitored cold chain). Biomarker measurement, including plasma p-tau217 and other dementia-relevant biomarkers, is performed in collaboration with the UK DRI Biomarker Factory, alongside standard clinical chemistry tests.
- Participants provide consent for recontact for future studies and trials and for linkage to NHS and other health-related records. External studies can request DTA to approach eligible participants via a controlled governance process; identifiable information is not transferred without consent.
- DTA sits within a rapidly developing UK dementia trial-readiness landscape and is designed to be interoperable with wider initiatives
- Sponsors and investigators of RCTs and clinical studies are invited to contact the DTA team to use the DTA services.

## Data Availability

All data produced in the present study are available upon reasonable request to the Dementia Trials Accelerator

## Authorship

### Funding

The Dementia Trials Accelerator, a join initiative of HDRUK and UKDRI, received funding from the Dame Barbara Windsor Dementia Goals program through the UK Medical Research Council.

### Authorship statement

WW wrote the first draft of the manuscript. All authors contributed to the design and delivery of the DTA, and critically reviewed the manuscript for content.

